# Prevalence and risk factors for long COVID after mild disease: a longitudinal study with a symptomatic control group

**DOI:** 10.1101/2022.09.15.22279958

**Authors:** Ana Beatriz C Cazé, Thiago Cerqueira-Silva, Adriele Pinheiro Bomfim, Gisley Lima de Souza, Amanda Canário Andrade Azevedo, Michelle Queiroz Aguiar Brasil, Nara Rúbia Santos, Ricardo Khouri, Jennifer Dan, Antonio Carlos Bandeira, Luciano Pamplona de Goes Cavalcanti, Manoel Barral-Netto, Aldina Maria Prado Barral, Cynara Gomes Barbosa, Viviane Sampaio Boaventura

**Author notes:** Corresponding author Viviane Sampaio Boaventura, Email address, Postal address: 121 Waldemar Falcão Street, Candeal, Salvador, Bahia, Brazil, CEP 40296. Those authors equally supervised this study (Cynara Gomes Barbosa, Viviane Sampaio Boaventura).

## Abstract

**Background:** There is limited data on the prevalence and risk factors for long COVID, with a shortage of prospective studies with appropriate control groups and adequate sample size. We therefore performed a prospective study to determine the prevalence and risk factors for long COVID.

**Methods:** We recruited patients age ≥ 15 years who were clinically suspected of having acute SARS-CoV-2 infection from September 2020 to April 2021. Nasopharyngeal swabs were collected for RT-PCR 3-5 days post symptom onset. Clinical and sociodemographic characteristics were collected using structured questionnaires from persons positive and negative for SARS-COV-2. Follow-up was performed by telephone interview to assess early outcomes and persistent symptoms. For COVID-19 cases, 5D-3L EuroQol questionnaire was used to assess the impact of symptoms on quality of life.

**Results:** We followed 814 participants (412 COVD-19 positive and 402 COVID-19 negative persons) of whom the majority (741 / 814) had mild symptoms. Both the COVID-19 positive and the COVID-19 negative groups had similar sociodemographic and clinical characteristics, except for the rate of hospitalization (15.8% vs 1.5%, respectively). One month after disease onset, 122 (29.6%) individuals reported residual symptoms in the COVID-19 positive group or the long COVID group versus 24 (6%) individuals in the COVID-19 negative group. In the long COVID group, fatigue, olfactory disorder, and myalgia were the most frequent symptoms which occurred in the acute phase. Compared to recovered patients, female sex, older age and having > 5 symptoms during the acute phase were risk factors for long COVID. Quality of life was evaluated in 102 out of 122 cases of long COVID with 57 (55.9%) reporting an impact in at least one dimension of the EuroQol 5D-3L questionnaire.

**Conclusion:** In this prospective study consisting predominantly of patients with mild disease, the persistence of symptoms after acute disease was highly associated with long COVID-19 (29.6% vs 6% of COVID negative group). The risk factors for long COVID were older age, female sex, and polysymptomatic acute disease.

## BACKGROUND

Long COVID has received less attention from the scientific community than acute COVID-19, representing less than 2% of COVID-19 publications in PubMed Clinical Queries, August 31, 2022). Several important aspects remain undefined such as its prevalence. Long COVID has been reported to effect between 7.5% to 89% of patients [1, 2, 29, 30]. Most of these studies do not include well-matched controls which is important because long COVID-symptoms are nonspecific and may be attributed to other factors such as the physical and mental impact of having COVID-19 infection or hospitalization [3,4]. Prospective studies enrolling symptomatic individuals with and without confirmed COVID-19 are necessary to better estimate the prevalence of long COVID and its associated risk factors.

Long COVID is anticipated to have a large impact on the healthcare system [5]. Identifying risk factors for developing long COVID will help mitigate its impact. Some risk factors have been suggested for long COVID; however, these studies consisted of previously hospitalized patients [1] or were retrospective analyses from electronic health records of not hospitalized patients (31). We therefore performed a longitudinal study of patients with COVID-like symptoms who tested positive or negative for SARS-CoV-2 to assess the prevalence and risk factors for long COVID and its impact on the quality of life.

## METHODS

### Study population

We recruited individuals age ≥ 15 years with clinical suspicion of acute SARS-CoV-2 infection in health care units from three municipalities of Bahia State-Brazil (Irecê, Campo Formoso, and Lauro de Freitas). Individuals who had difficulty in reporting symptoms, those who were > 20 days post symptom onset at the time of recruitment, or those who reported SARS-CoV-2 re-infection were excluded. The study was conducted from September 2020 to April 2021. The ancestral strain or gamma variant of SARS-CoV-2 was dominant in Brazil from November 2020 to August 2021 (**Additional file 1: Fig S1-2**) [6].

### Exposure measurement

All patients were tested for COVID-19 by nasopharyngeal RT-PCR. Only individuals who had an RT-PCR in the first ten days of disease onset were considered eligible for the follow-up. The Strengthening the Reporting of Observational Studies in Epidemiology (STROBE) protocol was followed.

### Recruitment strategy and data collected

Consecutive in-person recruitment included the application of a structured survey (Questionnaire 1) using Research Electronic Data Capture (REDCap) software hosted by the Gonçalo Moniz Institute (IGM/FIOCRUZ) in Bahia, Brazil. The following information was collected: sociodemographic characteristics (age, gender, skin color, body mass index, anthropometric data, municipality of residence), comorbidities (hypertension, diabetes, heart disease, chronic lung disease, other), date of symptom onset, acute symptoms (fever, myalgias, arthralgias, cough, shortness of breath, nasal congestion, sneezing, coryza, sore throat, sinus pain, retroocular pain, ear pain, headache, chest pain, chest discomfort, rash, abdominal pain, loss of appetite, vomiting, diarrhea (2 or more episodes in the last 24 hours), weakness, fatigue, lightheadedness or fainting, loss of smell, and loss of taste).

### Follow-up and outcome measurement

SARS-CoV-2 positive (COVID-19) and negative (non-COVID-19) patients who reported COVID-like symptoms were followed. Negative cases were recruited on the same day as positive cases before nasopharyngeal RT-PCR. The date of symptom onset was considered Day zero. A second structured questionnaire (Questionnaire 2) was conducted 30 to 60 days after disease onset by telephone to document the RT-PCR result, acute symptoms, and clinical outcome. Participants were classified as non-hospitalized or hospitalized in the intensive care unit (ICU) or non-ICU). To avoid memory bias, questionnaires 1 and 2 were applied in a short interval of time.

A third structured questionnaire was conducted 60 to 250 days after disease onset by telephone to evaluate residual symptoms (Questionnaire 3). Questions included residual or new symptoms: cough, fatigue, shortness of breath, headache, chest pain, dysphonia, dysphagia, loss of appetite, loss of smell, loss of taste, myalgias, arthralgias, fever, and other non-listed symptoms. Patients reporting suspected or confirmed re-infection were excluded from the final analysis. A schematic diagram of the study is provided (**Additional file 1**: **Fig S3**).

### Outcomes of interest

Long COVID was defined according to the Centers for Disease Control and Prevention classification [7]. Persistence of symptoms was defined as the presence of at least one residual symptom > 1 month post disease onset. The list of residual symptoms presented on the questionnaire was based on literature [8,9].

The European Quality of Life 5 Dimensions 3 Level (EQ-5D-3L) was conducted via telephone to assess health dimensions: mobility, self-care, daily activities, pain/discomfort and anxiety/depression. The EQ-visual analogue scale (VAS) was also conducted with 0 rated as worst health imaginable and 100 as the best imaginable health [10].

### Statistical analysis

Categorical data were described using absolute values and percentages/frequencies. Continuous data were described using mean (standard deviation, SD) and median (interquartile range, IQR). The correlation matrix was constructed based on the frequency of persistent symptoms reported by the patients. Multivariable logistic regression was performed to explore risk factors related to long COVID, applying the odds ratios (OR; 95% CI). Analyses were done in R (version 4.2.0) [26].

## RESULTS

In total, 1,268 patients with suspected SARS-CoV-2 infection were recruited and answered the first questionnaire 4 days (range of 3-5 days) post symptom onset. After excluding cases outside the RT-PCR recommended window period, reported re-infection, or those lost to follow-up, 814 (64.2%) patients were considered eligible for the analysis of residual symptoms (**Fig. 1, Additional file 1: Fig. S3**). Questionnaires 2 and 3 were conducted 40 (31-59) days and 102 (63-150) days post symptom onset, respectively. No difference in sociodemographic data and frequency of positive RT-PCR were observed between the included (n=814) and excluded (n=454) participants **(Additional file 1: Table S1**). Among the included participants, 412 tested positive (COVID-19 group) and 402 tested negative (non-COVID-19 group) for SARS-CoV-2. Demographic characteristics for the COVID and non-COVID groups were as follows, respectively: a median age of 36 (IQR) and 34 (IQR) years, female gender of 55% and 64%, mild disease in 84% and 99% (**Table 1**). More patients were hospitalized in the COVID-19 group compared to the non-COVID-19 group (15.9% vs 1.5%, respectively, **Table 1**). Follow-up time was 3.3 months for the COVID group and 3.7 months for the non-COVID group (**Additional file 1: Fig. S4**).

**Table 1:**
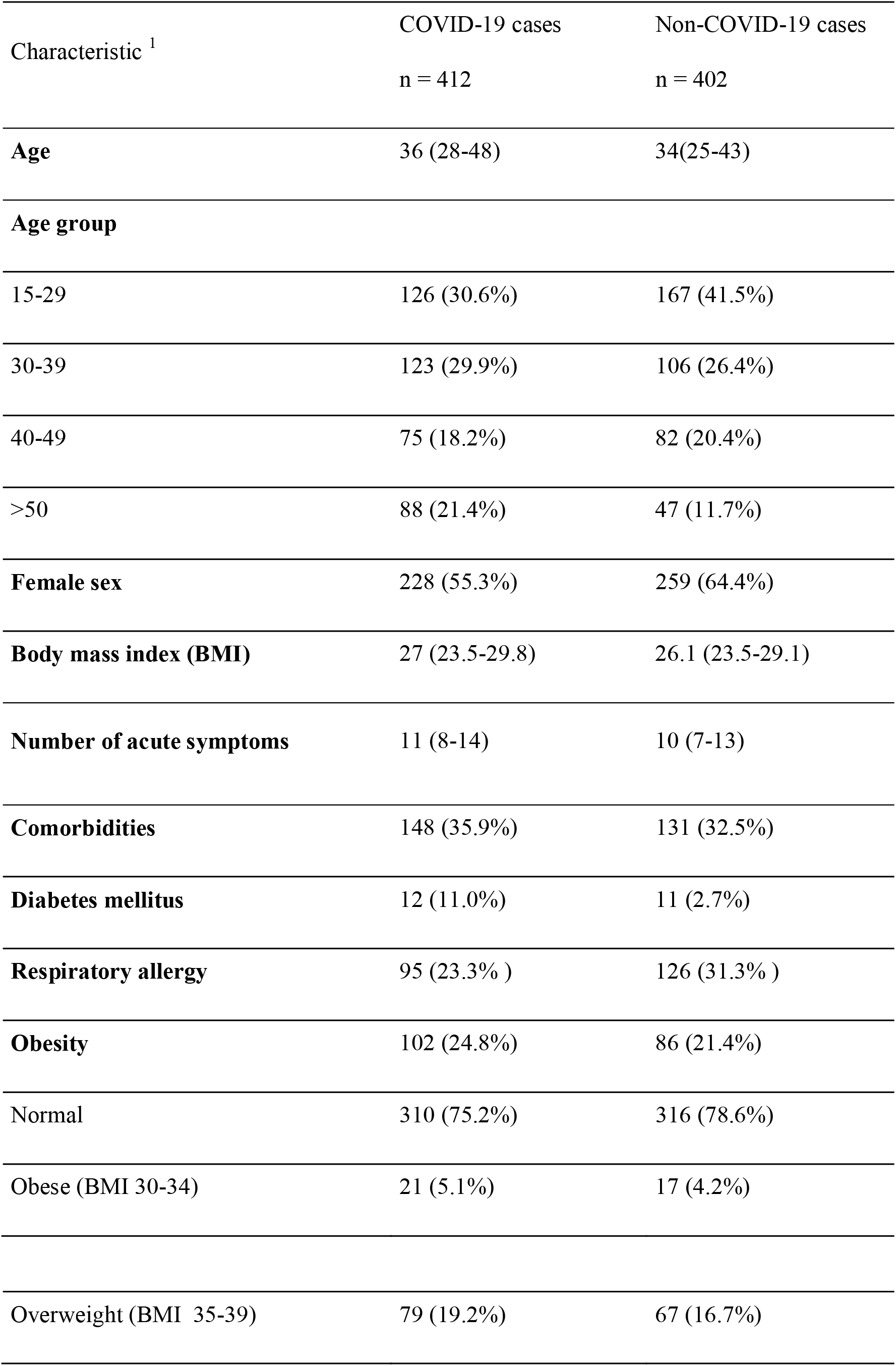
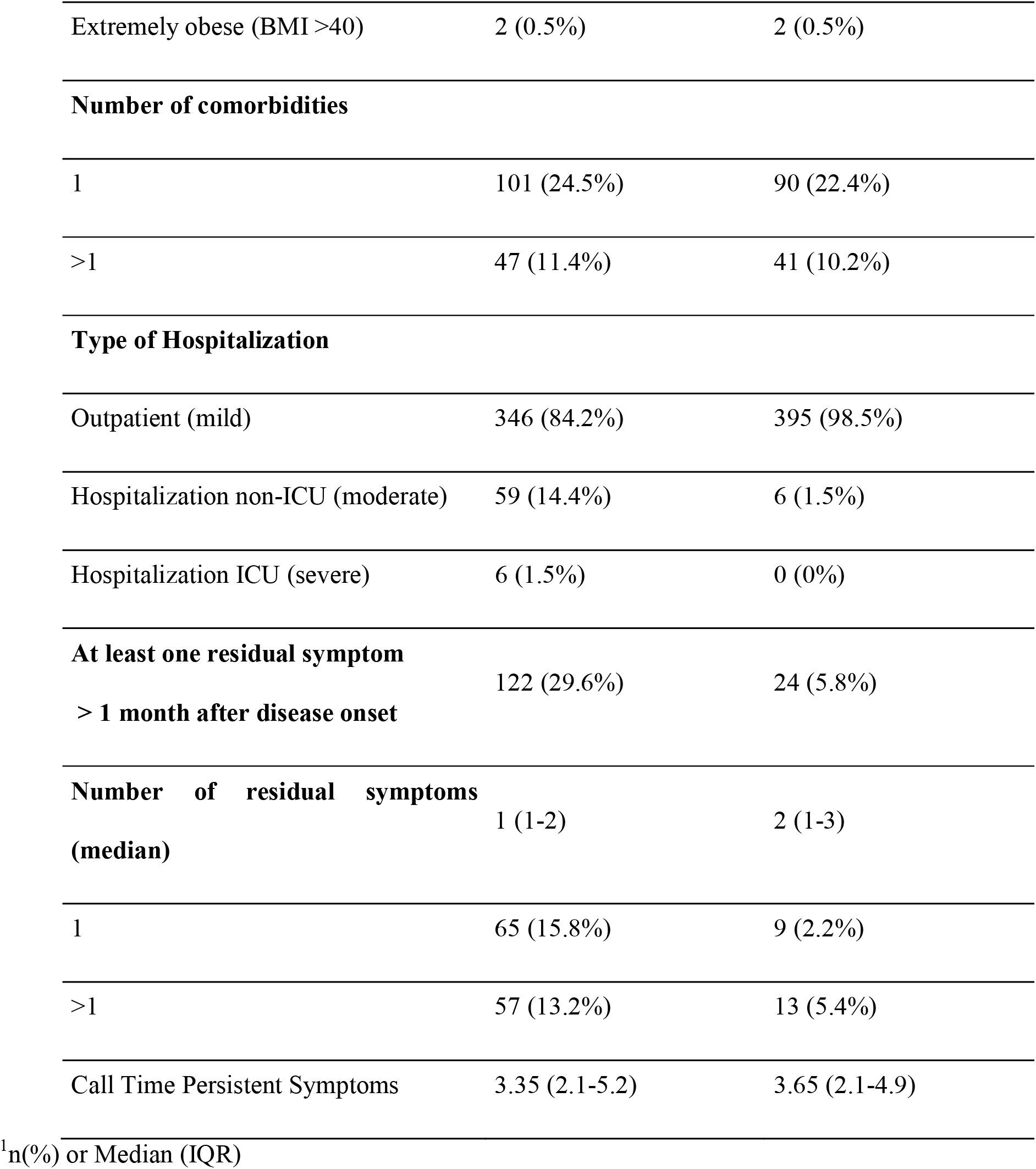
Clinical, sociodemographic characteristics and frequency of residual symptoms for COVID-19 and non-COVID-19 cases.

**Fig 1.**
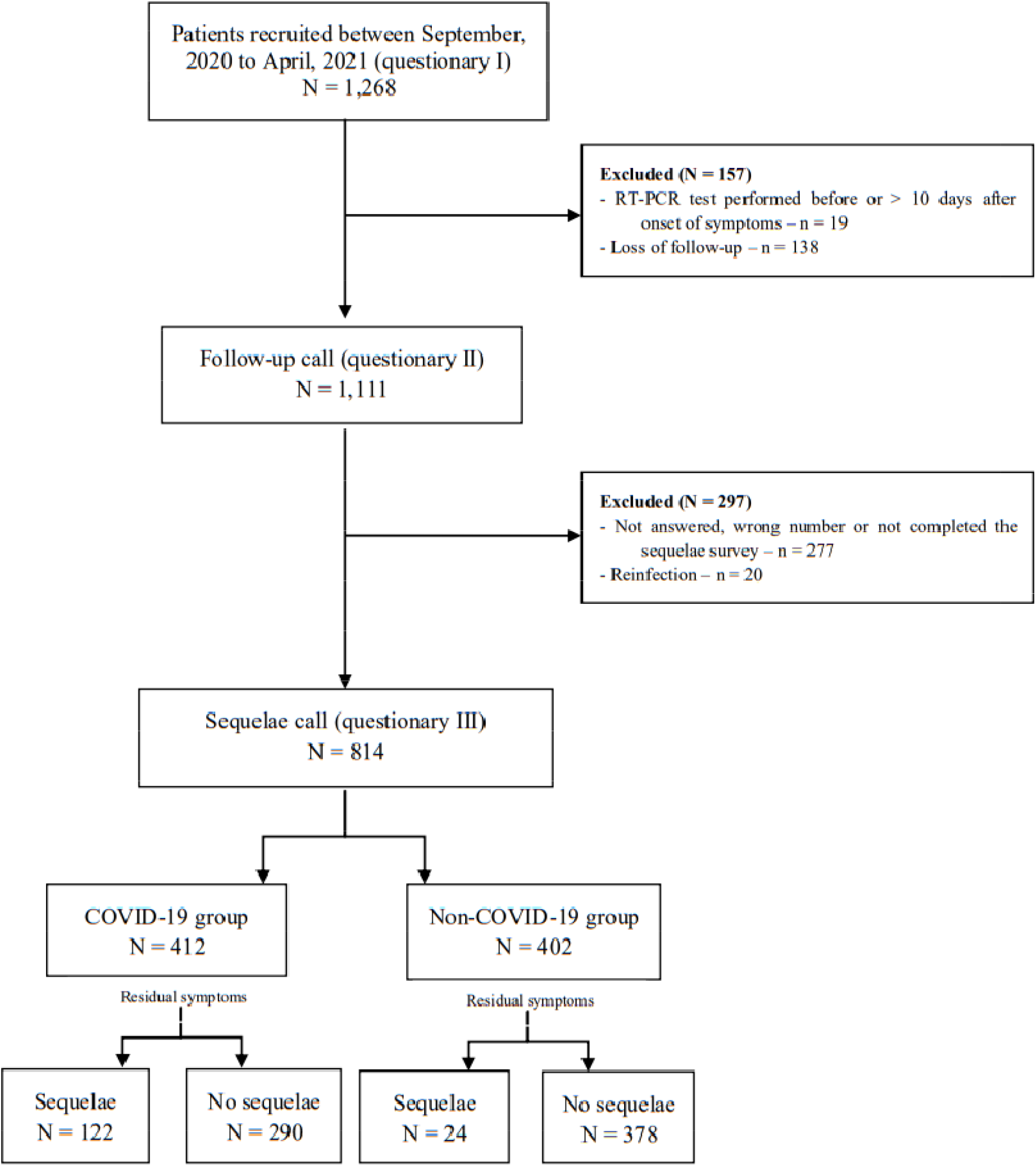
Flowchart of the study population.

After one month post disease onset, per the CDC definition for long COVID (September 16, 2021), 122/412 (29.6%) individuals from the COVID group reported residual symptoms and were classified as long COVID cases, while the remaining 290 individuals were fully recovered. From the non-COVID group, residual symptoms were reported by only 24 (6%) individuals (**Table 1**). Among long COVID cases, 57 (13.2%) presented with more than two residual symptoms.

Within the COVID group, fatigue (56, 13.7%), olfactory disorder (42, 9.9%), myalgia (36, 8.7%), gustatory disorder (27, 6.6%), and headache (26, 6.1%) were the residual symptoms most frequently reported (**Fig. 2**). These symptoms started mainly during the acute phase. Other symptoms not reported during the acute phase, such as memory loss and hair loss, were reported by the patients with long COVID (**Additional file 1: Table S2**). Compared to those who recovered, the long COVID group had more women (63.1% vs. 52.1%) and older individuals [40 (32-51) vs. 35 (28-47) years] (**Table 2**). Hospitalization for acute disease was comparable between long COVID (21/122, 17.2%) and recovered patients (44/290, 15.2%).

**Table 2:** Clinical, sociodemographic characteristics and frequency of residual symptoms for COVID cases with or without sequelae.

| Characteristic <sup>1</sup> | No Sequelae<br>n=290 | Sequelae<br>n=122 |
| --- | --- | --- |
| **Age** | 35 (28, 47) | 40 (32, 51) |
| **Age group** |  |  |
| 15-29 | 99 (34.1%) | 27 (22.1%) |
| 30-39 | 85 (29.3%) | 38 (31.1%) |
| 40-49 | 51 (17.9%) | 24 (19.7%) |
| >50 | 55 (19.0%) | 33 (27.0%) |
| **Female sex** | 151 (52.1%) | 77 (63.1%) |
| **BMI** | 26.9 (23.5, 29.7) | 27.2 (23.8, 30.1) |
| **Number of acute symptoms** | 11 (7, 14) | 12 (9, 15) |
| **Comorbidities** | 101 (34.8%) | 47 (38.5%) |
| **Diabetes mellitus** | 6 (2.1%) | 6 (2.1%) |
| **Respiratory allergy** | 63 (21.9%) | 32 (26.2%) |
| **Obesity** | 70 (24.1%) | 32 (26.2%) |
| Obese (BMI 30-34) | 12 (4.1%) | 9 (7.4%) |
| Overweight (BMI 35-39) | 57 (19.7%) | 22 (18.0%) |
| Extremely obese (BMI >40) | 1 (0.3%) | 1 (0.3%) |
| **Number of comorbidities** |  |  |
| 1 | 75 (25.9%) | 26 (21.3%) |
| >1 | 26 (9.0%) | 21 (17.2%) |
| **Time of disease onset** | 3.30 (2.00, 5.10) | 3.75 (2.12, 5.55) |
| **Hospitalization** | 44 (15.2%) | 21 (17.2%) |
| 1. n(%) or Median (IQR) |  |  |

**Fig. 2:**
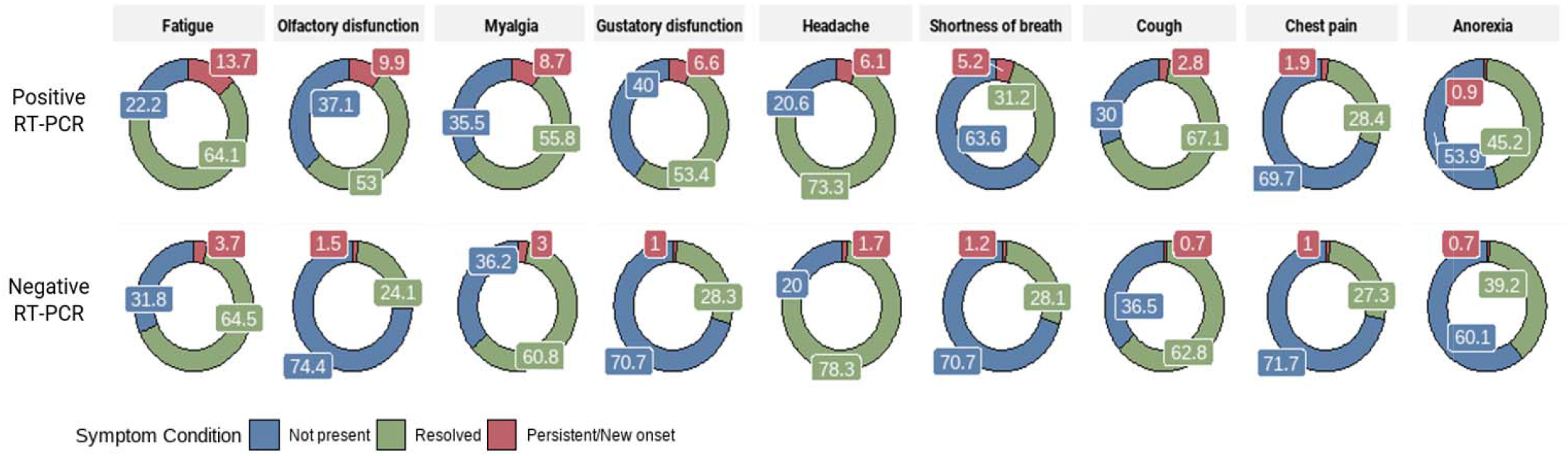
Frequency of acute and residual symptoms for COVID-19 and non-COVID-19 cases.

The risk of long COVID increased with age and was significant for persons 50 years of age or older (OR 2.44, 95% CI 1.29-4.66). Individuals with > five symptoms in the acute phase (OR 3.15, 95% CI 1.37-8.55) and females (OR 1.55, 95% CI 0.99-2.44) were more likely to develop long COVID (**Table 3**).

**Table 3.** Multivariable analysis of odds ratio (lower and upper confidence intervals) for presenting long COVID symptoms.

|  | OR | LCI* | UCI* | P value |
| --- | --- | --- | --- | --- |
| **Age group (reference 14-29 years)** |  |  |  |  |
| 30-39 | 1.71 | 0.95 | 3.12 | 0.078 |
| 40-49 | 2.04 | 1.02 | 4.10 | 0.045 |
| ≥ 50 | 2.44 | 1.29 | 4.66 | 0.006 |
| **Female sex** | 1.55 | 0.99 | 2.44 | 0.055 |
| **> 5 acute symptoms** | 3.15 | 1.37 | 8.55 | 0.012 |

\*LCI and UCI - lower and upper confidence intervals, respectively

Co-occurrence of long COVID symptoms was also observed. Fatigue was reported by at least 50% of those that complained of gustatory dysfunction, anorexia, dysphonia, chest pain, headache, breathlessness, or myalgia. Moreover, olfactory dysfunction was reported by 80% of participants with gustatory dysfunction (**Additional file 1: Fig. S5**).

Finally, we have evaluated the impact of long COVID on the quality of life. The EQ-5D-3L questionnaire was administered to 102 out of 122 patients. Fifty-seven patients (55.8%) reported alteration in at least one dimension of QoL, and 41 patients (41.1%) had an impact in the dimension of pain and depression (**Additional file 1: Table S3**).

## DISCUSSION

We have followed RT-PCR positive and negative patients with COVID-like symptoms from the first days post symptom onset to 63-150 days post symptom onset to determine the prevalence of long COVID-19. Long COVID occurred in almost one-third of test-positive cases, the majority with mild acute disease, compared to 6% of negative patients with residual symptoms, supporting the association between SARS-CoV-2 infection and residual symptoms. The risk of long COVID increased with age > 50 years old, female gender, and the development of > five symptoms during the acute phase. Together these findings demonstrate that residual symptoms after mild disease were mainly associated with COVID-19 with well-defined demographic and clinical risk factors.

The proportion of patients with COVID-19 who developed long COVID has varied from 7.5% to 89% [1, 2, 28, 29]. Most studies only included patients who tested positive for SARS-CoV-2. One strength of our study is the inclusion of a comparator group of symptomatic patients, recruited in the same time/location, who tested negative by SARS-CoV-2 RT-PCR. Additionally, most symptoms investigated in the follow-up questionnaires were evaluated at the time of recruitment, in a blinded manner, prior to the SARS-CoV-2 test result. The inclusion of a control group is important considering that many residual symptoms attributed to long COVID, such as fatigue and headache, are nonspecific and could be triggered or aggravated by stress and psychosocial problems connected to the global health crisis [11-13]. Indeed, we observed that non-confirmed COVID-19 cases reported similar residual symptoms after acute illness, albeit at a much lower frequency. Few studies have included a control group composed of symptomatic patients that tested negative for SARS-CoV-2 [14, 30]. In a cohort of healthcare workers, olfactory disorders and hair loss were related to long COVID while positive and negative cases both had similar frequencies of exhaustion/burnout and fatigue [14]. In a population-based digital retrospective cohort including outpatients and well-matched controls, 62 symptoms were associated with long COVID [30]. The inclusion of a control group is even more valuable for hospitalized cases, considering the increased risk of sequelae associated with iatrogenic procedures for treating severe disease [15]. Together, these findings highlight the importance of including a control group when determining the frequency of sequelae to avoid overestimating long COVID cases.

We observed a similar proportion of long COVID in groups with mild and moderate/severe disease [16,17]. We enrolled a small number of severe COVID-19 cases and therefore could not conclude an association between disease severity and sequelae. Severe COVID-19 increases the risk of long COVID by 1.2 to 8x among hospitalized cases compared to those not hospitalized [18]. Nonetheless, the finding that long COVID occurs in a significant proportion of individuals after mild disease highlights its potential impact on the healthcare system, as COVID-19 variants and sub-variants continue to spread worldwide.

Fatigue was the most frequent sequelae and occurred isolated or associated with other clinical manifestations. As previously reported [19], we also observed a vast impact of long COVID on the quality of life. Although the mechanism is unknown, an exacerbation of the immunological response and multisystemic involvement [20] seems to be involved. Studies investigating the pathogenesis of long COVID are essential to improve treatment and attenuate the impact.

Similar to previous studies [3, 9, 17, 21-23, 30], we found that long COVID was more prevalent in women and could be attributed to a greater propensity for women to go to a clinic [22]. Indeed, females represented 55% of the COVID group and 64% of the non-COVID group in our study. Sex hormone differences in immune response [24] and autoimmunity triggered by SARS-CoV-2 have been suggested to be involved in the disease pathogenesis of long COVID [23]. Other studies have found that individuals with more symptoms during the acute phase were at a higher risk for developing long COVID [9, 14]. There are conflicting results regarding age in the risk for developing long COVID. In a cohort of healthcare workers in Switzerland, the majority of whom had mild disease, young age was associated with a higher risk for developing long COVID [14], in contrast to our findings. Additional prospective studies including multicenter cohorts with larger sample sizes and older patients should be performed to evaluate if the risk for developing long COVID differs among age groups, racial groups, and ethnicities. Prospective studies to develop predictive models and algorithms should be performed for the early detection of long COVID cases.

Long COVID has different definitions. We have used the CDC’s definition for long COVID (September 16, 2021), that includes persistence of symptoms after four weeks of the onset of disease. Recently, a WHO-coordinated Delphi consensus established a period of more than three months after COVID-19 onset for the case definition of long COVID [25]. In our study, only 445 of the calls were performed three months after disease onset. By this definition, long COVID was detected in 68/216 (31.5%) of our patients. Residual symptoms were also observed in 17/229 (7.4%) of the non-COVID-19 patients. As observed in our study, the prevalence of long COVID may vary between studies in the absence of a universal consensus to define long COVID [4]. This impacts the estimation of the prevalence of long COVID cases worldwide.

This study has some limitations. The loss to follow-up was around 36% of patients and may have influenced the estimates of residual symptoms. Similar demographic and clinical characteristics were observed among patients that were included and the ones that were lost to follow-up. In addition, most participants were young, with mild disease, which does not permit generalizing these findings to the elderly and those with severe disease.

Moreover, the participants were aware of their diagnosis of COVID-19 when questionnaire 3 was applied, which may have led to an overestimation of persistent symptoms. Finally, the short time to follow-up did not allow for an analysis of the duration of long COVID.

## CONCLUSIONS

Long COVID was detected in 29.6% of patients with mild COVID-19 disease, with older age, female sex, and polysymptomatic acute disease as the main risk factors for persistent symptoms. Estimating the long COVID prevalence is important for preparing the healthcare system to assist and guide these patients. Further studies should be conducted to evaluate the impact of different variants of SARS-CoV-2 on long COVID and the potential impact of vaccines in reducing residual symptoms.

## Data Availability

Availability of data and materials: The dataset generated and/or analyzed during the current study are not publicly available due to legal and ethical restrictions. Dataset are available upon request.

## LIST OF ABBREVIATIONS

STROBE: Strengthening the Reporting of Observational Studies in Epidemiology
RT-PCR: Reverse Transcriptase Polymerase Chain Reaction
REDCap: Research Electronic Data Capture software
IGM/FIOCRUZ: Gonçalo Moniz Institute
ICU: Intensive Care Unit
SD: Standard Deviation
IQR: Interquartile Range

## DECLARATIONS

### Ethics approval and consent to participate

Informed consent was obtained from all participants. The Ethics in Research Committee of the Gonçalo Moniz Research Center approved this study (approval no. 4.315.319/202).

### Consent for publication

Not applicable

### Availability of data and materials

The dataset generated and/or analyzed during the current study are not publicly available due to legal and ethical restrictions. Dataset are available upon request.

### Authors’ contributions

A.B.C.C. and V.B structured the original draft. V.B. and C.B. were responsible for conceptualization and methodology. A.P.B., G.L.S, A.C.A.A, M.Q.A.B, N.R.S and A.B.C.C collected data from the patients. T.C.S made the formal analysis statistics. A.M.P.B. and V.B. participated in the project administration and in the funding acquisition. TCS, JD, ACB, LPGC, MBN, ABN, CGB substantively revised the manuscript. All authors read and approved the final manuscript.

## Acknowledgements

We wish to thank all patients who were willing to participate in the study. In addition, we would like to thank Hospital Aeroporto and Municipal Health Secretariats of Irecê and Campo Formoso for logistical support and to Roberta de Carvalho Freitas for patient-related support.

