## Supplementary material for "Prevalence and risk factors for long COVID after mild disease: a longitudinal study with a symptomatic control group"

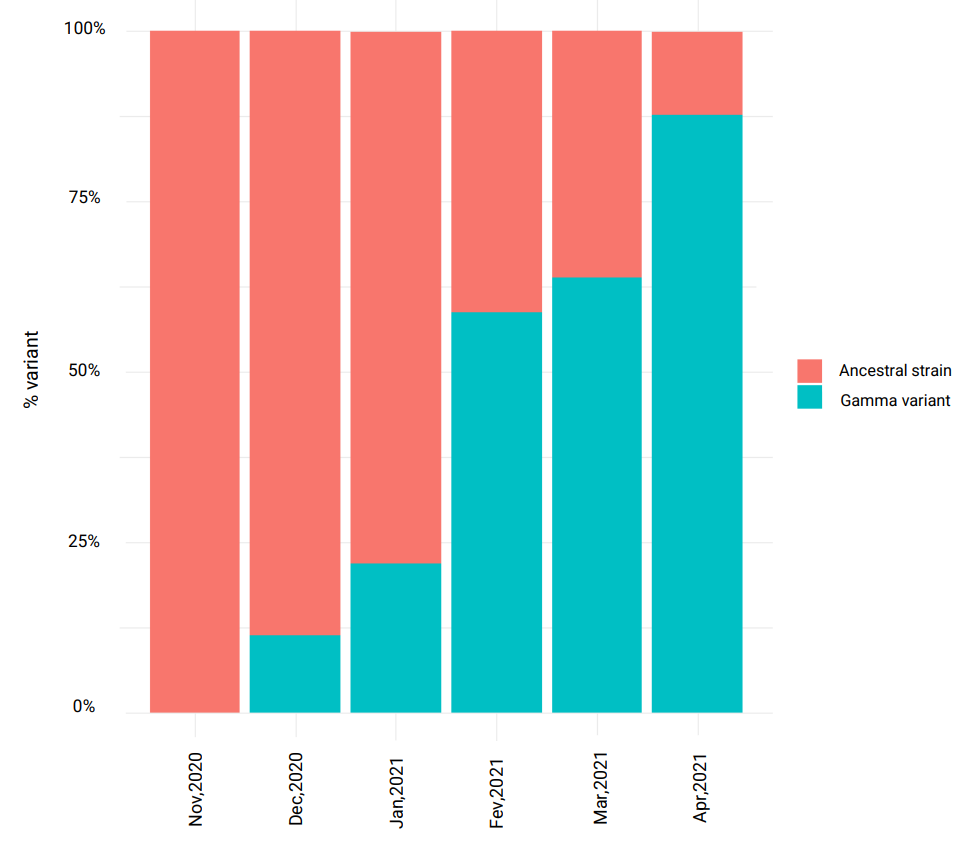
**Fig. S1** - Prevalence of SARS-CoV-2 variants of concern in Brazil from November, 2020, to April, 2021.

Data obtained from the Fiocruz Genomic Network [6].

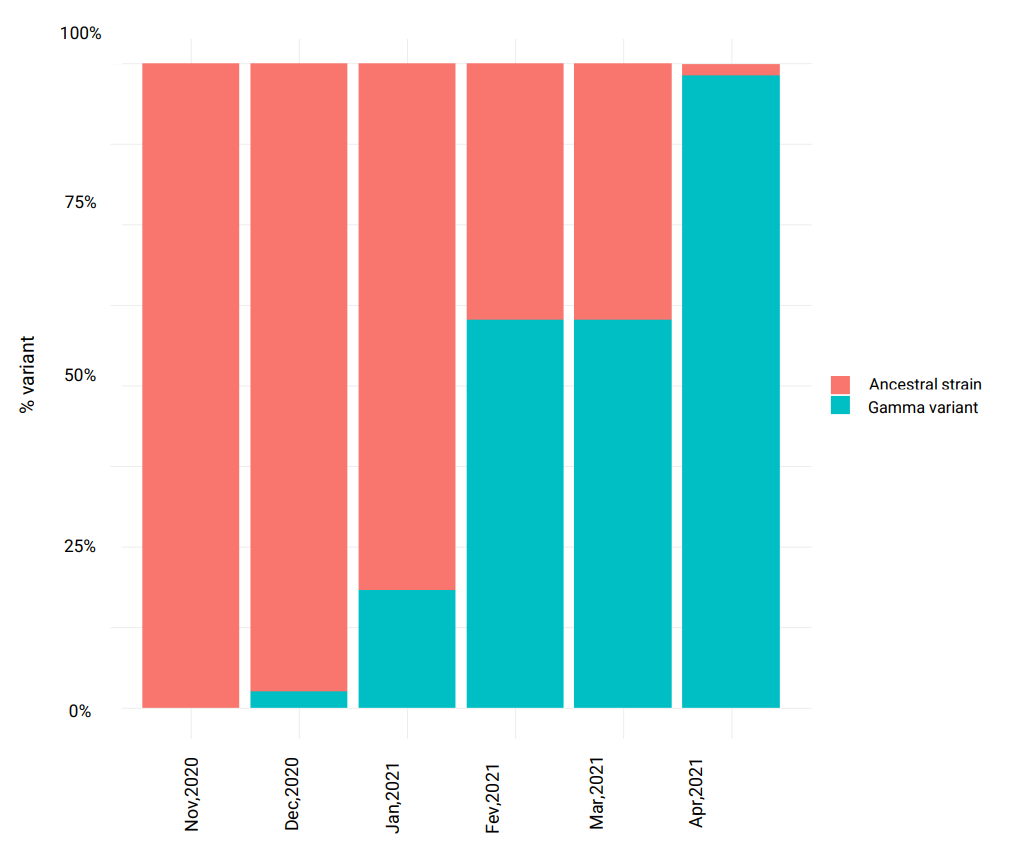
**Fig. S2** - Prevalence of SARS-CoV-2 variants of concern in Northeast, Brazil, from November, 2020 to April, 2021.

Data obtained from the Fiocruz Genomic Network [6].

**Fig. S3** – Diagram of the questionnaire application: time and data collected.
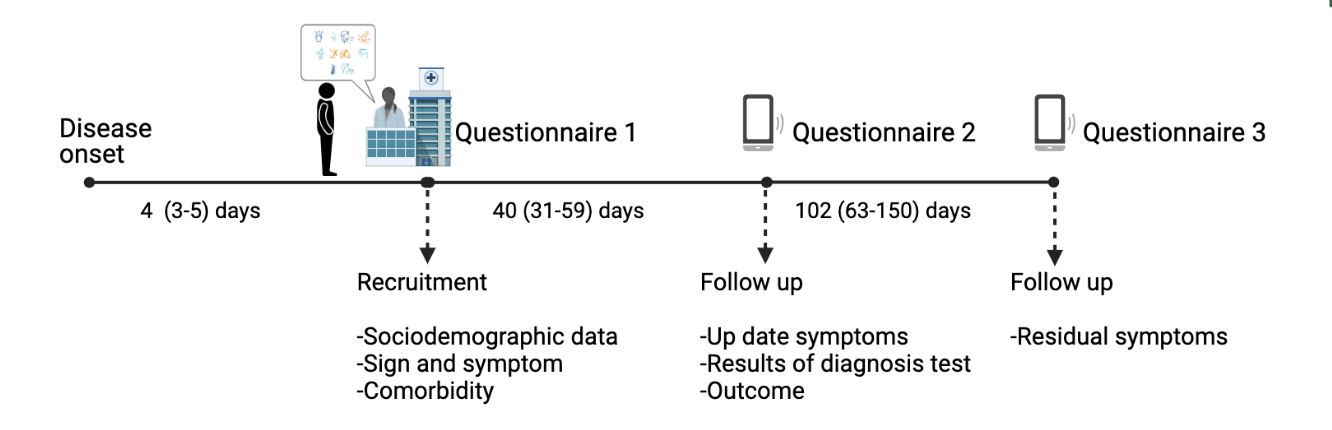

Figure created with BioRender [27].

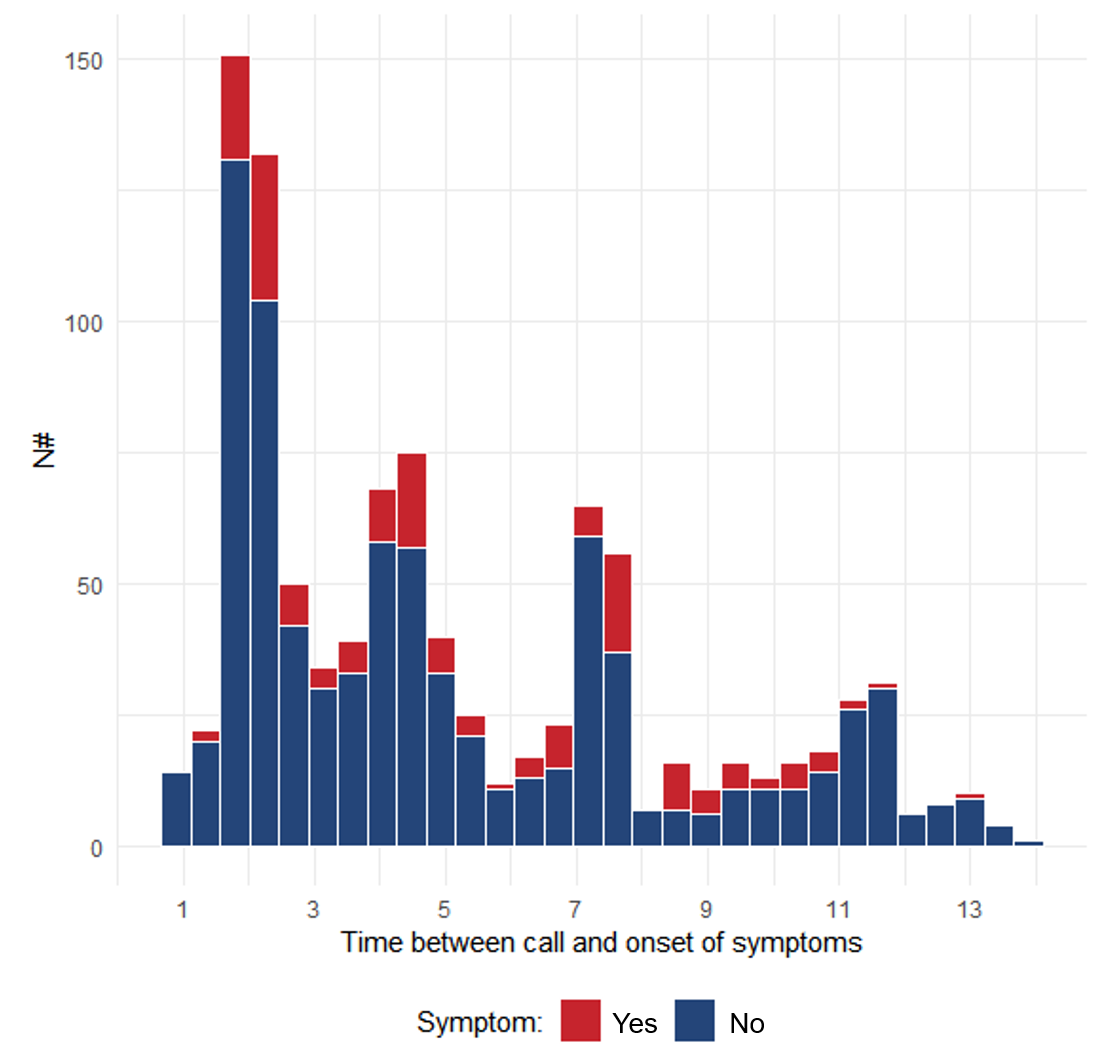
**Fig. S4** - Time between residual symptoms call (questionnaire 3) and onset of symptoms.

**Fig. S5** - Correlation matrix with the frequency of residual symptoms of long COVID patients.

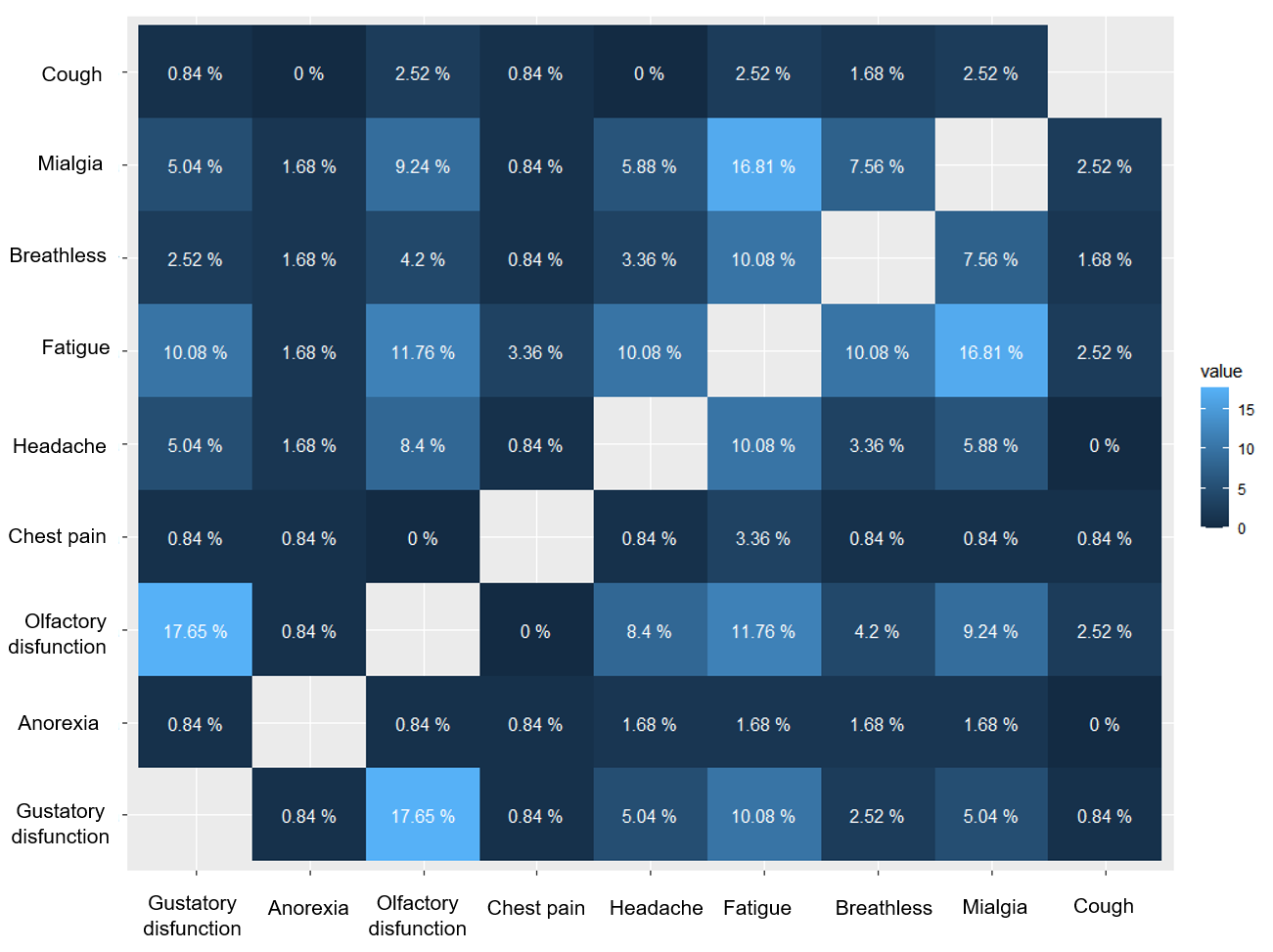

**Table S1** – Characteristics comparing loss of follow-up and included patients.

| Characteristic ^1^ | Loss of follow-up  n = 454 | Included patients  n = 814 |
| --- | --- | --- |
| **Age** | 37(28-47) | 35 (27-45) |
| **Female sex** | 271 (59.7%) | 487 (59.8%) |
| **Positive RT-PCR** | 239 (52.6%) | 412 (50.6%) |
| **BMI** | 26 (23.6-28.9) | 26.5 (23.5-29.4) |
| **Diabetes mellitus** | 23 (17.2%) | 23 (2.8%) |
| **Respiratory allergy** | 138 (30.6%) | 221 (27.1% ) |
| **Type of Hospitalization** |  |  |
| Outpatient (mild) | 437 (96.3%) | 741 (91.0%) |
| Hospitalization non-ICU (moderate) | 12 (2.6%) | 65 (8.0%) |
| Hospitalization ICU (severe) | 5 (1.1%) | 6 (0.7%) |
| 1. n(%) or Median (IQR) |  |  |

**Table S2** – Other symptoms reported by patients with long COVID.

| Other reported  residual symptoms | n (%) |
| --- | --- |
| Loss of memory | 15 (12.3%) |
| Hair loss | 9 (7.4%) |
| Joint pain | 4 (3.3%) |
| Ear pain / ear disorder | 4 (3.3%) |

**Table S3** - Assessment of quality of life using EuroQol instrument EQ-5D-3L.

| Quality of Life^1^ | Long COVID  n=102 |
| --- | --- |
| **Mobility** |  |
| I have no problems in walking about | 94 (92.1%) |
| I have some problems in walking about | 8 (7.8%) |
| I am confined to bed | 0 |
| **Self-care** |  |
| I have no problems with self-care | 96 (94.1%) |
| I have some problems washing or dressing myself | 6 (5.9%) |
| I am unable to wash or dress myself | 0 |
| **Usual activities** |  |
| I have no problems with performing my usual activities | 73 (71.6%) |
| I have some problems with performing my usual activities | 28 (27.5%) |
| I am unable to perform my usual activities | 1 (0.9%) |
| **Pain / discomfort** |  |
| I have no pain or discomfort | 60 (58.8%) |
| I have moderate pain or discomfort | 1 (0.9%) |
| I have extreme pain or discomfort | 41 (40.2%) |
| **Anxiety / depression** |  |
| I am not anxious or depressed | 60 (58.8%) |
| I am moderately anxious or depressed | 29 (28.4%) |
| I am extremely anxious or depressed | 13 (12.7%) |
| **Score global** | 80 (70,90) |
